# Changes in Use of HIV Prevention Methods in East Zimbabwe over the Course of the COVID-19 Pandemic: A Longitudinal Population Study

**DOI:** 10.1101/2025.10.21.25338467

**Authors:** S Bagnay, L Moorhouse, C Nyamukapa, B Tsenesa, M Skovdal, P Mandizvidza, B Moyo, R Morris, R Maswera, S Gregson

## Abstract

**Introduction:** COVID-19 posed substantial challenges for sustaining use of HIV prevention services in sub-Saharan African populations. However, data on changes in use of HIV primary prevention methods within the general population over the course of the pandemic remain scarce. We investigated these changes in east Zimbabwe.

**Methods:** Data were taken from three rounds of a large-scale open-cohort population survey conducted before (July 2018 to December 2019), during (February to July 2021), and after (July 2022 to March 2023) stringent COVID-19 control measures. Changes in use of HIV prevention methods, underlying sexual risk-behaviours for HIV acquisition, and socio-demographic determinants were measured using descriptive statistics and logistic regression analyses of serial cross-sectional and cohort data.

**Results:** In the serial cross-sectional analysis, in uninfected men, use of HIV prevention methods in non-regular sexual partnerships decreased from 40.4% (N=261) before COVID-19 to 35.3% during COVID-19 (AOR=0.79, 95%CI, 0.56-1.12; N=295), and 27.1% after COVID-19 (AOR=0.57, 0.39-0.83; N=293). This decrease reflected primarily a reduction in use of male condoms. No changes were found in overall use of HIV prevention methods in non-regular sexual partnerships in uninfected women. In women with sexual risk behaviours, PrEP use rose from 0.5% (N=1001) before COVID-19 to 3.2% during COVID-19 (AOR=6.07, 2.29-16.13; N=785), and 4.2% after COVID-19 (AOR=7.54, 2.93-19.41; N=928). 19.3%, 20.2% and 21.3% of sexually active men in the before, during and after COVID-19 surveys, respectively, reported a non-regular partnership in the last 12 months. The corresponding proportions for women were 8.3%, 7.2% and 7.1%. Similar results were found in the cohort analyses.

**Conclusion:** In east Zimbabwe, men engaging in non-regular partnerships had lower use of HIV prevention methods during and after the COVID-19 pandemic than before the pandemic despite continuing high levels of sexual risk behaviour. In women, increases in PrEP use partly compensated for lower use of other prevention methods. Epidemic preparedness plans should ensure that access to HIV prevention methods is maintained.

## 1. Introduction

The emergence of the COVID-19 pandemic in 2020 profoundly impacted the daily lives of individuals around the globe [1]. Governments enforced stringent containment and mitigation measures, including restrictions on movement and social mixing. While these measures were imperative to control the spread of SARS-CoV-2 infection, their impact extended beyond the pandemic with far-reaching consequences for other diseases including HIV and AIDS [2]. Research from sub-Saharan Africa (SSA) suggests that COVID-19 exacerbated pre-existing vulnerabilities and inequalities that drive local HIV epidemics [3, 4].

Previous public health emergencies have significantly impacted sexual and reproductive health (SRH). During the Ebola outbreak in West Africa in 2014-2015, a sharp decline in use of sexual health [5] and family planning services [6] was seen. Recovery to pre-outbreak levels was not achieved for several months, highlighting the detrimental impact of health emergencies on the progress made in improving SRH over recent decades [7].

Now that the emergency phase of the COVID-19 pandemic has passed, it is crucial to establish the effects of the pandemic and to take stock of its impact on local HIV epidemics in high-burden countries. We investigate changes in use of HIV prevention methods, underlying sexual risk behaviours (SRB), and their sociodemographic determinants in east Zimbabwe across different phases of the pandemic. Describing and understanding these changes will support efforts to prepare for future pandemics and to design and implement more resilient health systems in Zimbabwe and comparable SSA countries.

### 1.1 Use of HIV prevention methods in SSA during COVID-19

An early modelling study found that, if COVID-19 social distancing interventions did not reduce non-regular sexual partners, interruptions in condom supply and peer education services in 50% of people could increase new HIV infections by 1.19 times over a 1-year period [7]. In populations such as in east Zimbabwe, where HIV risk behaviour remained high [2], it is especially pertinent to establish whether reductions in use of prevention methods occurred that could have increased new HIV infections.

Empirical evidence suggests that restrictions and lockdowns in SSA countries led to temporary discontinuation or reduced operation of health programmes [3, 8–10]. These disruptions impacted negatively on use of HIV prevention methods in SSA through multiple pathways, including supply-side challenges, limited access to services [8–11], and exacerbation of pre-existing funding cuts, with further resources being diverted away from HIV prevention during COVID-19 [3]. Individual-level factors, such as fear of SARS-CoV-2 infection when visiting clinics or shops [4, 10, 12] or the presence of police roadblocks limiting mobility [13] also discouraged individuals from seeking prevention services. Temporal changes in these response measures and fears might have affected the demand for, availability of, access to, and ultimately use of HIV prevention methods amongst those at risk of acquiring HIV across the different stages of the pandemic. A recent systematic review found a growing body of research on the impact of COVID-19 on local HIV epidemics. However, most studies in SSA were from early in the pandemic and focused on HIV testing and use of antiretroviral therapy (ART) [10]. Data on the impact of COVID-19 on use of primary HIV prevention methods within the general population is particularly limited.

Here, we use data from three rounds of an open-cohort population survey in Manicaland, east Zimbabwe, covering periods shortly before, during and immediately after COVID-19. Through a combination of serial cross-sectional and cohort analyses, we address the following objectives:

1. To describe changes in the sociodemographic characteristics and social behaviours of HIV-negative individuals aged 15-54 in east Zimbabwe over the course of the COVID-19 pandemic;
2. To describe changes in SRBs for HIV acquisition among sexually-active HIV-negative adults in the general population; and
3. To describe changes in use of HIV prevention methods among sexually-active HIV-negative adults and in those in the priority population for HIV prevention (i.e. those with SRBs).

Throughout the paper, when we refer to the ‘during’ COVID-19 period, we mean the period when the most stringent containment and mitigation measures were implemented to control the spread and adverse health impacts of successive variants of SARS-CoV-2. The ‘after’ COVID-19 period refers to the time when these measures were relaxed, and governments and populations sought to return to normal. We recognise that COVID-19 was still considered a Public Health Emergency of International Concern [14] during this latter period.

## 2. Methods

### 2.1 Data

Data were taken from rounds 7-9 of the Manicaland General Population Open-Cohort Survey (Manicaland Survey), covering eight study sites representing four main socioeconomic strata in Manicaland (urban, peri-urban, estates, and rural areas). The Manicaland survey gathers a broad range of information on sociodemographic characteristics, marital history, SRBs, and use of HIV prevention methods. For information on the data collection procedures and sampling scheme, please refer to Table 1, the studies by Morris et al. [2] and Gregson et al. [15], and the Manicaland Centre website (http://www.manicalandhivproject.org/).

**Table 1.** Data collection procedures for the Manicaland HIV/STD Prevention Project general population open cohort survey conducted in East Zimbabwe.

| Study round | Data collection | Interview method | Provider-initiated HIV testing and dried blood spot samples available |
| --- | --- | --- | --- |
| Round 7<br>'before' COVID-19 | July 2018 to December 2019 | Face-to-face interviews | Yes |
| Round 8<br>'during' COVID-19 | February 2021 to July 2021 | Telephone interviews | No* |
| Round 9<br>'after' COVID-19 | July 2022 to March 2023 | Face-to-face interviews (household census) & telephone interviews (individual survey) | No* |

\*HIV status measured by self-reported questions.

The current study used data for HIV-negative participants aged 15-54 years who self-reported not using Treatment-as-Prevention (TasP) due to the possibility of being HIV-positive. Two types of statistical analyses were conducted: serial cross-sectional and longitudinal cohort analyses. The former included all eligible HIV-negative individuals who participated in each study round; the latter was restricted to HIV-negative individuals who participated in all three survey rounds. This combined approach sought to provide insights into both population-level trends and individual changes over time, offering a more comprehensive understanding of the outcomes while acknowledging that differences in findings could arise.

### 2.2 Study measures

#### Sociodemographic characteristics and social behaviours

Socioeconomic status (SES) was measured in quintiles [16]. Marital status was coded into four categories: never married, married or cohabiting, divorced or separated, and widowed. Dichotomised variables were constructed for three social behaviours that can influence engagement in SRBs and may be affected by COVID-19 or associated control measures: (1) regular visits to bars and beerhalls, (2) drug use, and (3) whether women experienced intimate partner violence (IPV), consisting of sexual violence, physical violence, or both.

#### The priority population and sexual risk behaviour(s)

The priority population includes HIV-negative sexually-active participants who self-report engaging in ≥1 of five SRBs in the past year, and, therefore, would benefit from using HIV prevention methods. The SRBs were also used to establish HIV risk categories, similar to the categorisation developed by Morris et al. [2] (Table 2).

**Table 2.** Summary of study variables measuring sexual risk behaviours and HIV risk categories, Manicaland, east Zimbabwe.

| Study participants | Sexual risk behaviours | HIV risk categories |
| --- | --- | --- |
| HIV-negative sexually-active participants aged 15-54 | - | Low risk |
|  | (1) Multiple sexual partners in the past 12 months | Medium risk |
|  | (2) Concurrent sexual partners |  |
|  | (3) Transactional sex with any of the last three sexual partners in the past month |  |
|  | (4) One or more non-regular sexual partner(s) in the past 12 months | High risk |
|  | (5) Age-disparate relationship (10+ years) with any of the last three sexual partners | - |

#### Use of HIV prevention methods

VMMC use was defined as self-reporting full medical circumcision ever (“ever had VMMC”) or in the last 12 months (“recent VMMC”). PrEP use was defined as using PrEP as a current HIV prevention method. Male and female condom use were measured by self-reported condom use with regular or non-regular sexual partners in the past two weeks. Sexual abstinence and faithfulness for HIV prevention were defined as self-reporting current engagement in these practices for HIV prevention.

Summary variables measuring use of ≥1 effective method(s) of HIV prevention were derived using the above variables on VMMC (males only), PrEP, and male and female condom use among those reporting non-regular sexual partners.

### 2.3 Statistical analysis

The survey data were analysed and measured for differences between: (1) Round 8 (during COVID-19) *versus* Round 7 (before COVID-19), and (2) Round 9 (after COVID-19) *versus* Round 7 (before COVID-19), in serial cross-sectional and cohort analyses. In Round 7, young women aged 15-24 years and young men aged 15-29 years were over-sampled due to intervention trials being conducted within these groups [17]. Consequently, all younger individuals were eligible to participate, and data from this round were weighted to account for over-sampling.

To meet the study objectives, sociodemographic characteristics, social behaviours (HIV-negative participants), SRBs, and use of HIV prevention methods (HIV-negative sexually-active participants) were described using proportions and 95% confidence intervals (95% CI). Logistic regression analysis was used to measure changes and compare exposure across each category in sociodemographic profile, social behaviours, SRBs for HIV acquisition, and use of prevention methods in both analyses. Adjusted Odds Ratios (AOR), 95% Confidence Intervals (95% CI), and *p*-values were calculated. Odds ratios were adjusted for age and site type, with site type adjusted for age. A *p*-value of ≤0.05 was considered statistically significant. Some odds ratios could not be calculated due to low statistical power. Forest plots were generated to visualise and compare differences in SRB (RStudio version 2025.05.1) and use of HIV prevention methods across rounds. Sankey diagrams were created to visualise the flow of participants across HIV risk categories in the cohort analysis (Microsoft PowerBI Sankey Diagram visual).

All analyses were disaggregated by sex (male/female) and conducted in STATA version 17.0. Due to space limitations in the main manuscript, additional materials, including cohort tables, are available in an open-access repository on Zenodo [18].

## 3. Results

9803 (91.7%) participants completed the individual questionnaire in round 7, 8497 (86.0%) in round 8, and 9,618 (84.0%) in round 9. Those outside the age-range, who were HIV-positive, had an unknown HIV status, or used TasP were excluded from the analysis. Those who seroconverted between survey rounds were excluded from the cohort analyses. Table 3 summarises the study participant numbers.

**Table 3.** Participant breakdown per study round, sex and analysis type in Manicaland, Zimbabwe.

|  | **Males (n)** | **Females (n)** |
| --- | --- | --- |
| *Round 7 – serial cross-sectional analysis* | 3181 | 4135 |
| *Round 8 – serial cross-sectional analysis* | 2770 | 3398 |
| *Round 9 – serial cross-sectional analysis* | 3254 | 3981 |
| *Cohort analysis\** | 668 | 1186 |

\*Participants taking part in all three study rounds.

### 3.1 Changes in sociodemographic characteristics and social behaviours of HIV-negative individuals over the course of COVID-19

#### Sociodemographic characteristics

Table 4 compares the sociodemographic characteristics of the study population across the phases of COVID-19 in a serial cross-sectional analysis. In both sexes, the study population was older during and after COVID-19 than before COVID-19. For males, the proportions living in urban and peri-urban areas were higher during and after COVID-19 than before COVID-19, while the proportions residing in estate and rural sites were reduced. For females, during COVID-19, the proportion living in peri-urban areas increased and the proportion in estates reduced; however, after the pandemic, these differences disappeared, and the proportions living in urban and rural sites increased and decreased, respectively. No statistically significant differences in site type were found in the cohort analysis; outcomes are available through an open-access repository [18]. After adjusting for age and site type, in both sexes, the study population became more concentrated in the central (3^rd^ poorest) quintile during and after COVID-19, with fewer people in the poorest and least poor quintiles – a trend also observed in the cohort. Fewer women were married during and after COVID-19 than before COVID-19. In the cohort, this reduction only became statistically significant after COVID-19 [18].

**Table 4.** Comparisons of sociodemographic characteristics and social behaviours in HIV-negative adults aged 15-54 in Manicaland (Zimbabwe) after and during COVID-19 *versus* before the pandemic in a serial cross-sectional analysis.

| Socio-demographic characteristics and social behaviours | MALES |  |  |  |  |  |  |  | FEMALES |  |  |  |  |  |  |  |
| --- | --- | --- | --- | --- | --- | --- | --- | --- | --- | --- | --- | --- | --- | --- | --- | --- |
|  | After COVID-19 |  |  | During COVID-19 |  |  | Before COVID-19 |  | After COVID-19 |  |  | During COVID-19 |  |  | Before COVID-19 |  |
|  | n=3254 |  |  | n=2770 |  |  | n=3181 |  | n=3981 |  |  | n=3398 |  |  | n=4135 |  |
|  | After vs before COVID-19 |  |  | During vs before COVID-19 |  |  |  |  | After vs before COVID-19 |  |  | During vs before COVID-19 |  |  |  |  |
|  | AOR [95% CI]<sup>a,c</sup> | % | n | AOR [95% CI]<sup>a,c</sup> | % | n | %<sup>b</sup> | n | AOR [95% CI]<sup>a,c</sup> | % | n | AOR [95% CI]<sup>a,c</sup> | % | n | %<sup>b</sup> | n |
| *Sociodemographic characteristics* |  |  |  |  |  |  |  |  |  |  |  |  |  |  |  |  |
| *Age groups* |  |  |  |  |  |  |  |  |  |  |  |  |  |  |  |  |
| 15-19 | 0.72 [0.65-0.80] * | 24.8 | 808 | 0.76 [0.69-0.84] * | 25.9 | 716 | 26.4 | 986 | 0.66 [0.60-0.73] * | 19.5 | 777 | 0.73 [0.67-0.80] * | 21.2 | 717 | 21.5 | 1112 |
| 20-24 | 0.78 [0.68-0.88] * | 17.1 | 557 | 0.74 [0.65-0.84] * | 16.2 | 448 | 17.6 | 656 | 0.67 [0.60-0.74] * | 17.5 | 696 | 0.62 [0.56-0.69] * | 16.0 | 541 | 18.8 | 969 |
| 25-34 | 0.95 [0.85-1.06] | 22.8 | 741 | 0.96 [0.86-1.06] | 22.7 | 628 | 24.0 | 744 | 1.16 [1.06-1.27] * | 25.7 | 1023 | 1.24 [1.14-1.35] * | 27.1 | 917 | 27.5 | 948 |
| 35-44 | 1.24 [1.13-1.36] * | 21.2 | 690 | 1.26 [1.15-1.37] * | 21.8 | 604 | 20.2 | 502 | 1.44 [1.34-1.56] * | 22.9 | 910 | 1.46 [1.36-1.57] * | 22.2 | 752 | 20.2 | 694 |
| 45-54 | 1.68 [1.47-1.92] * | 14.1 | 458 | 1.57 [1.39-1.78] * | 13.5 | 374 | 11.8 | 293 | 1.57 [1.41-1.76] * | 14.4 | 575 | 1.45 [1.31-1.61] * | 13.6 | 462 | 12.0 | 412 |
| *Site type* |  |  |  |  |  |  |  |  |  |  |  |  |  |  |  |  |
| Urban | 2.08 [1.86-2.32] * | 26.2 | 854 | 1.52 [1.36-1.70] * | 20.5 | 569 | 14.5 | 480 | 1.43 [1.31-1.57] * | 24.9 | 991 | 1.04 [0.95-1.13] | 19.2 | 652 | 18.7 | 805 |
| Peri-urban | 1.12 [1.02-1.24] * | 25.1 | 816 | 1.28 [1.17-1.41] * | 27.9 | 772 | 23.1 | 727 | 0.98 [0.90-1.06] | 27.6 | 1100 | 1.17 [1.09-1.26] * | 31.4 | 1063 | 28.1 | 1151 |
| Estates | 0.62 [0.57-0.68] * | 23.1 | 752 | 0.69 [0.63-0.75] * | 24.8 | 687 | 32.6 | 1018 | 1.01 [0.93-1.10] | 22.2 | 885 | 0.87 [0.80-0.94] * | 19.6 | 665 | 21.9 | 905 |
| Rural | 0.80 [0.73-0.88] * | 25.6 | 832 | 0.86 [0.78-0.94] * | 26.8 | 742 | 29.9 | 956 | 0.74 [0.69-0.80] * | 25.2 | 1005 | 0.93 [0.87-1.00] | 29.8 | 1009 | 31.2 | 1274 |
| *Socioeconomic status quintile* |  |  |  |  |  |  |  |  |  |  |  |  |  |  |  |  |
| Poorest | 0.46 [0.37-0.57] * | 4.4 | 144 | 0.55 [0.45-0.68] * | 5.6 | 155 | 9.8 | 305 | 0.56 [0.47-0.67] * | 4.6 | 184 | 0.73 [0.62-0.86] * | 6.8 | 230 | 99.0 | 368 |
| 2<sup>nd</sup> poorest | 1.08 [0.97-1.20] | 41.3 | 1342 | 0.98 [0.89-1.09] | 42.2 | 1168 | 46.5 | 1488 | 1.15 [1.05-1.26] * | 40.1 | 1597 | 1.13 [1.04-1.24] * | 41.9 | 1421 | 40.3 | 1661 |
| 3<sup>rd</sup> poorest | 1.22 [1.09-1.37] * | 28.4 | 922 | 1.39 [1.24-1.56] * | 29.2 | 808 | 22.3 | 705 | 1.13 [1.03-1.25] * | 27.7 | 1104 | 1.19 [1.08-1.31] * | 27.4 | 928 | 24.0 | 999 |
| 4<sup>th</sup> poorest | 0.98 [0.86-1.11] | 24.8 | 806 | 0.91 [0.80-1.03] | 22.3 | 618 | 20.1 | 642 | 0.90 [0.81-1.00] | 26.0 | 1033 | 0.81 [0.73-0.91] * | 22.8 | 774 | 25.0 | 1037 |
| Least poor | 0.88 [0.57-1.36] | 1.2 | 38 | 0.52 [0.32-1.85] * | 0.8 | 21 | 1.2 | 41 | 0.87 [0.62-1.23] | 1.6 | 62 | 0.61 [0.41-0.91] * | 1.0 | 36 | 1.7 | 69 |
| *Marital status* |  |  |  |  |  |  |  |  |  |  |  |  |  |  |  |  |
| Never married | 1.10 [0.96-1.25] | 44.4 | 1444 | 1.03 [0.90-1.17] | 43.4 | 1203 | 43.7 | 1599 | 1.15 [1.03-1.29] * | 23.9 | 952 | 1.18 [1.06-1.32] * | 24.6 | 834 | 23.8 | 1178 |
| Married | 0.92 [0.81-1.04] | 51.9 | 1688 | 1.01 [0.89-1.14] | 53.3 | 1476 | 52.5 | 1479 | 0.79 [0.72-0.87] * | 61.6 | 2452 | 0.84 [0.77-0.92] * | 62.5 | 2117 | 64.8 | 2527 |
| Divorced | 0.97 [0.73-1.29] | 3.4 | 109 | 0.85 [0.63-1.15] | 2.9 | 81 | 3.5 | 97 | 1.29 [1.11-1.48] * | 10.9 | 435 | 1.12 [0.97-1.29] | 9.7 | 329 | 8.6 | 332 |
| Widowed | 1.59 [0.65-3.87] | 0.4 | 13 | 1.39 [0.54-3.56] | 0.4 | 10 | 0.2 | 6 | 1.22 [0.96-1.54] | 3.6 | 142 | 1.12 [0.89-1.40] | 3.2 | 109 | 2.8 | 98 |
| *Social behaviours* |  |  |  |  |  |  |  |  |  |  |  |  |  |  |  |  |
| *Visited bars or beerhalls at least once last month* |  |  |  |  |  |  |  |  |  |  |  |  |  |  |  |  |
| Yes | 0.49 [0.44-0.55] * | 18.3 | 595 | 0.46 [0.40-0.51] * | 16.9 | 468 | 31.0 | 931 | 0.96 [0.58-1.60] | 0.7 | 26 | 0.92 [0.54-1.59] | 0.6 | 21 | 0.7 | 27 |
| No | - | 81.7 | 2659 | - | 83.1 | 2302 | 69.0 | 2250 | - | 99.3 | 3955 | - | 99.4 | 3368 | 99.3 | 4108 |
| *Currently takes drugs for pleasure* |  |  |  |  |  |  |  |  |  |  |  |  |  |  |  |  |
| Yes | 0.15 [0.10-0.22] * | 0.8 | 25 | 0.59 [0.45-0.78] * | 2.9 | 81 | 4.8 | 150 | 0.24 [0.05-1.04] | 0.1 | 2 | 1.84 [0.74-4.56] | 0.4 | 12 | 0.2 | 8 |
| No | - | 99.2 | 3229 | - | 97.1 | 2689 | 95.2 | 3031 | - | 99.9 | 3979 | - | 99.6 | 3377 | 99.8 | 4127 |

| Experienced intimate partner violence |  |  |  |  |  |  |  |  |  |  |  |  |  |  |  |  |
| --- | --- | --- | --- | --- | --- | --- | --- | --- | --- | --- | --- | --- | --- | --- | --- | --- |
| Yes | - | - | - | - | - | - | - | - | 0.31 [0.26-0.37] * | 4.5 | 177 | 0.52 [0.44-0.60] * | 7.0 | 237 | 12.2 | 504 |
| No | - | - | - | - | - | - | - | - | - | 95.5 | 3804 | - | 93.0 | 3152 | 87.8 | 3631 |

<sup>a</sup> Odds Ratios for age are adjusted for site type, odds ratios for site type are adjusted for age, and all other variables are adjusted for age and site type.
<sup>b</sup> Percentages for the before COVID-19 period are weighted to account for oversampling of young people.
\* Asterisks are used to highlight statistically significant differences (*p*-value ≤0.05).
<sup>c</sup> All regression analyses account for clustering.

#### Social behaviours

In both analyses, men had lower odds of visiting bars and beerhalls and reporting recreational drug use during and after COVID-19 than before COVID-19 (Table 4). Fewer women than men reported these practices, with no statistically significant differences across survey rounds. Women’s reports of IPV decreased between rounds, from 12.2% before COVID-19 to 4.5% and 4.3% at the end of the pandemic in the serial cross-sectional and cohort analyses, respectively.

### 3.2 Changes in sexual risk behaviours in the general population and in the priority population for HIV prevention

Percentages of males reporting having started sex were close to 70% across all survey periods in the serial cross-sectional analysis [18]. Females had lower adjusted odds of reporting sexual initiation during COVID-19 [78.6%; AOR=0.72, 0.64-0.80] and after COVID-19 [80.6%; AOR=0.84, 0.74-0.94] than before COVID-19 [81.2%]. Within the cohort, the proportion of males reporting having started sex increased progressively from 79.1% before COVID-19 to 84.4% after COVID-19 [AOR=2.15, 1.54-3.02] [18]. For females, there was a non-statistically significant decrease during COVID-19.

In sexually active males, the only statistically significant change was a reduction in concurrent sexual partnerships after COVID-19 compared to before COVID-19 in the serial cross-sectional analysis [5.2% *versus* 6.6%; AOR=0.76, 0.59-0.98] (Figure 1b; [18]). In the same analysis, an increasing trend was found in non-regular partnerships from 19.3% before COVID-19 to 20.2% during COVID-19 [AOR=1.07, 0.92-1.26] and 21.3% after COVID-19 [AOR=1.16, 1.00-1.36] [18]. For females, both analyses showed reductions in multiple sexual partnerships within the last 12 months during [serial cross-sectional: 3.5% *versus* 6.1%; AOR=0.57, 0.44-0.72; cohort: 2.1% *versus* 4.0%; AOR=0.51, 0.30-0.87] and after COVID-19 [serial cross-sectional: 2.9% *versus* 6.1%; AOR=0.47, 0.37-0.60; cohort: 1.4% *versus* 4.0%; AOR=0.35, 0.19-0.64], compared to before COVID-19 (Figure 1c-d/g-h; [18]). A similar pattern was found for transactional sex. However, no changes were found in non-regular partnerships in the last 12 months [serial cross-sectional: 7.2% during COVID-19 *versus* 8.3% before COVID-19, AOR=0.94, 0.78-1.14; 7.1% after COVID-19 *versus* 8.3% before COVID-19, AOR=0.91, 0.76-1.09] or in concurrent partnerships or age-disparate partnerships [18].

**Figure 1.**
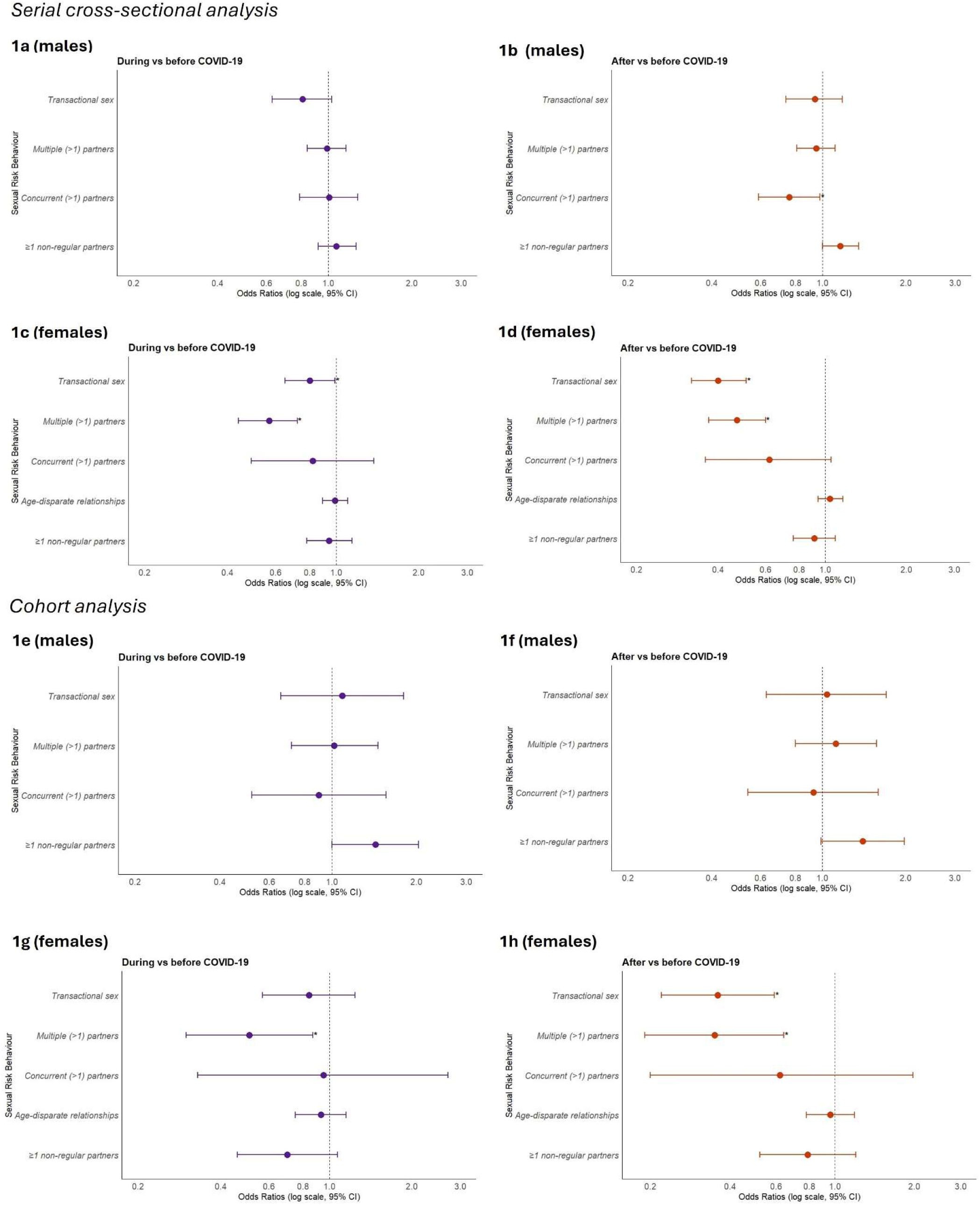
Forest plots (a-h) illustrating adjusted odds ratios and 95% confidence intervals for selected sexual behaviours during different phases of COVID-19 in east Zimbabwe. Serial cross-sectional and cohort analyses. Asterisks (*) highlight statistically significant differences (*p*-value ≤0.05).

No changes in proportions in HIV risk categories were observed between the before and during COVID-19 periods in the serial cross-sectional analysis [18]. However, males were less likely to be categorised as low-risk after COVID-19 [AOR=0.85, 0.74-0.97] than before COVID-19, while more females were categorised as low-risk [AOR=1.52, 1.30-1.78]. Males in the cohort were more likely to report high-risk behaviour during [AOR=1.58, 1.13-2.20] and after COVID-19 [AOR=1.71, 1.23-2.38] than before COVID-19 [18]. Females in the cohort had lower odds of being in the medium-risk group after COVID-19 than before COVID-19 [AOR=0.34, 0.19-0.60]. Figure 2 shows a Sankey diagram illustrating the flows between HIV risk categories for the cohort participants. Many individuals moved between categories, yet overall proportions of males and females in these categories changed only slightly between survey periods.

**Figure 2.**
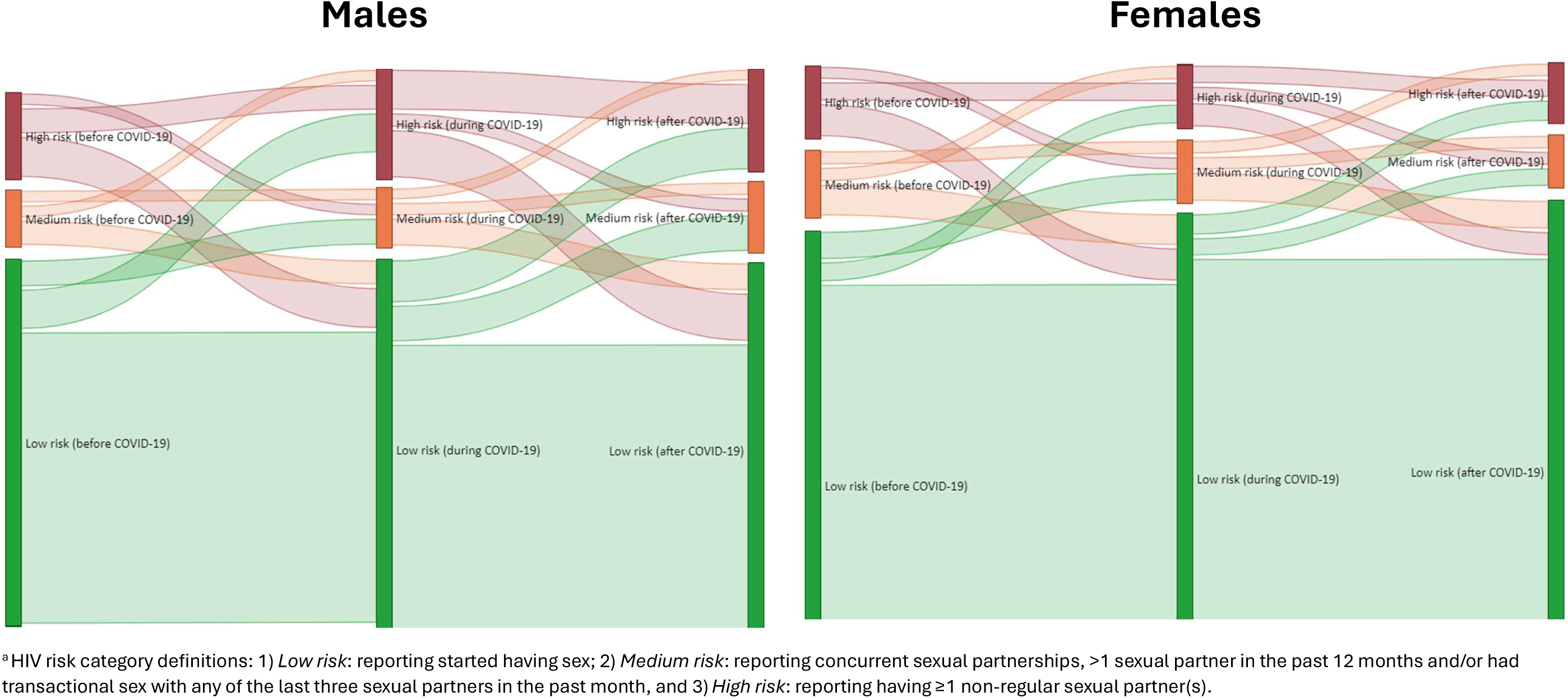
Sankey diagrams summarising changes in the proportions of individuals reporting sexual risk behaviours for HIV infection in a cohort of HIV-negative adults aged 15-54 years at baseline (before COVID-19) followed over three study rounds - before, during, and after COVID-19 in Manicaland, Zimbabwe^a^.

The odds of males being in the priority population for HIV prevention methods increased between the before COVID-19 and after COVID-19 periods in the serial cross-sectional analysis [AOR=1.18, 1.03-1.35] and in the cohort analysis [AOR=1.41, 1.05-1.90] [18]. For females, no statistically significant differences were found.

### 3.3 Changes in use of HIV prevention methods in sexually-active HIV-negative adults and in the priority population

In the serial cross-sectional analysis, in uninfected men, use of ≥1 effective method of HIV prevention (i.e. PrEP, male condoms or female condoms) in non-regular sexual partnerships decreased from 40.4% before COVID-19 to 35.3% during COVID-19 [AOR=0.79, 0.56-1.12], and 27.1% after COVID-19 [AOR=0.57, 0.39-0.83] (Table 5; Figure 3). No changes in use of ≥1 HIV prevention method in non-regular sexual partnerships were found for uninfected women. Similar results were found in the cohort analysis (Figure 3; [18]).

**Figure 3.**
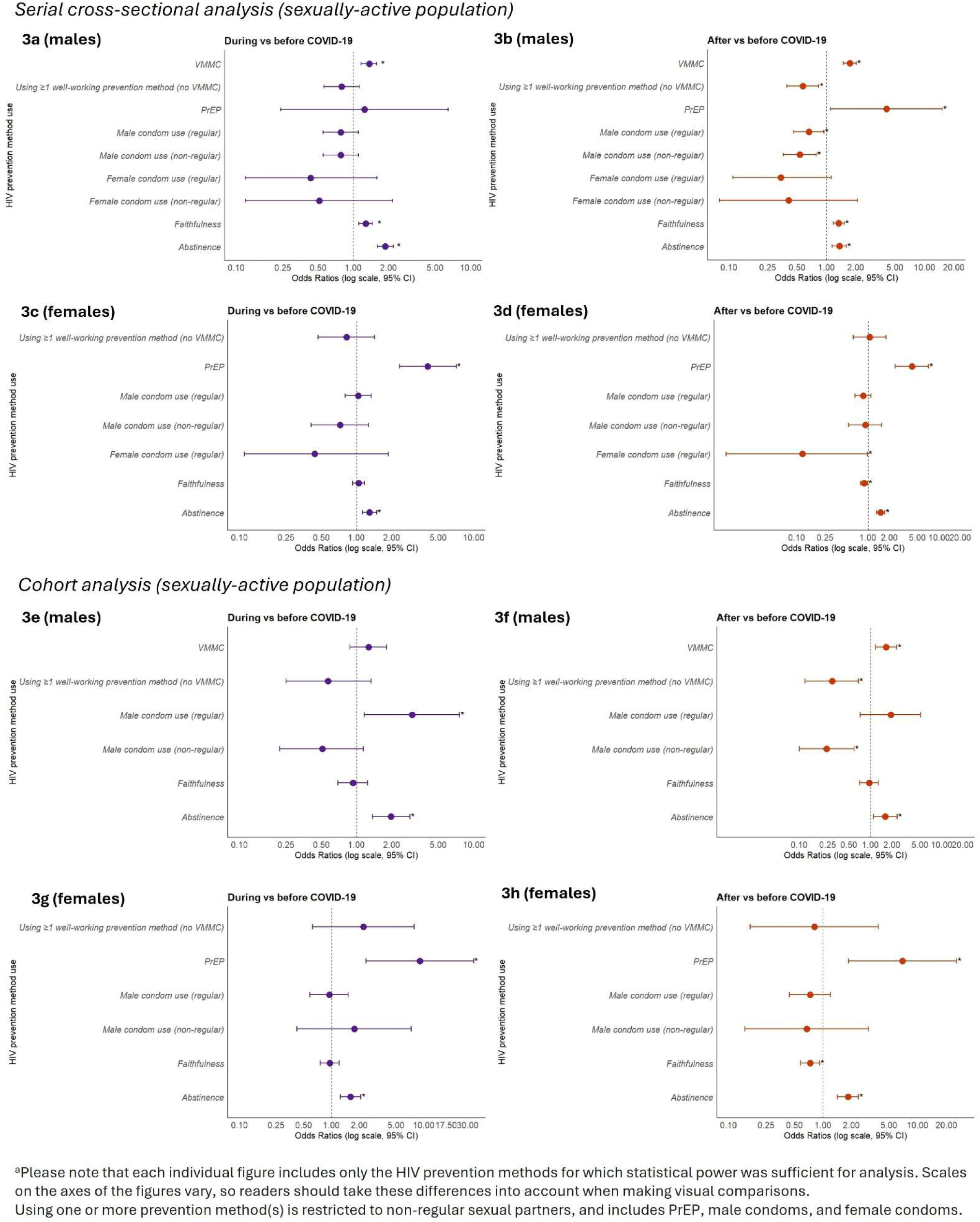
Log-transformed Forest Plots (a-h) illustrating HIV prevention method use among males and females, comparing after and during COVID-19 to before COVID-19, using data from a general population cohort in east Zimbabwe. Serial cross-sectional and cohort analyses^a^. Odds Ratios are adjusted for age- and site-type. Significant differences are marked with asterisks (*) for *p*-value ≤0.05.

**Table 5.**
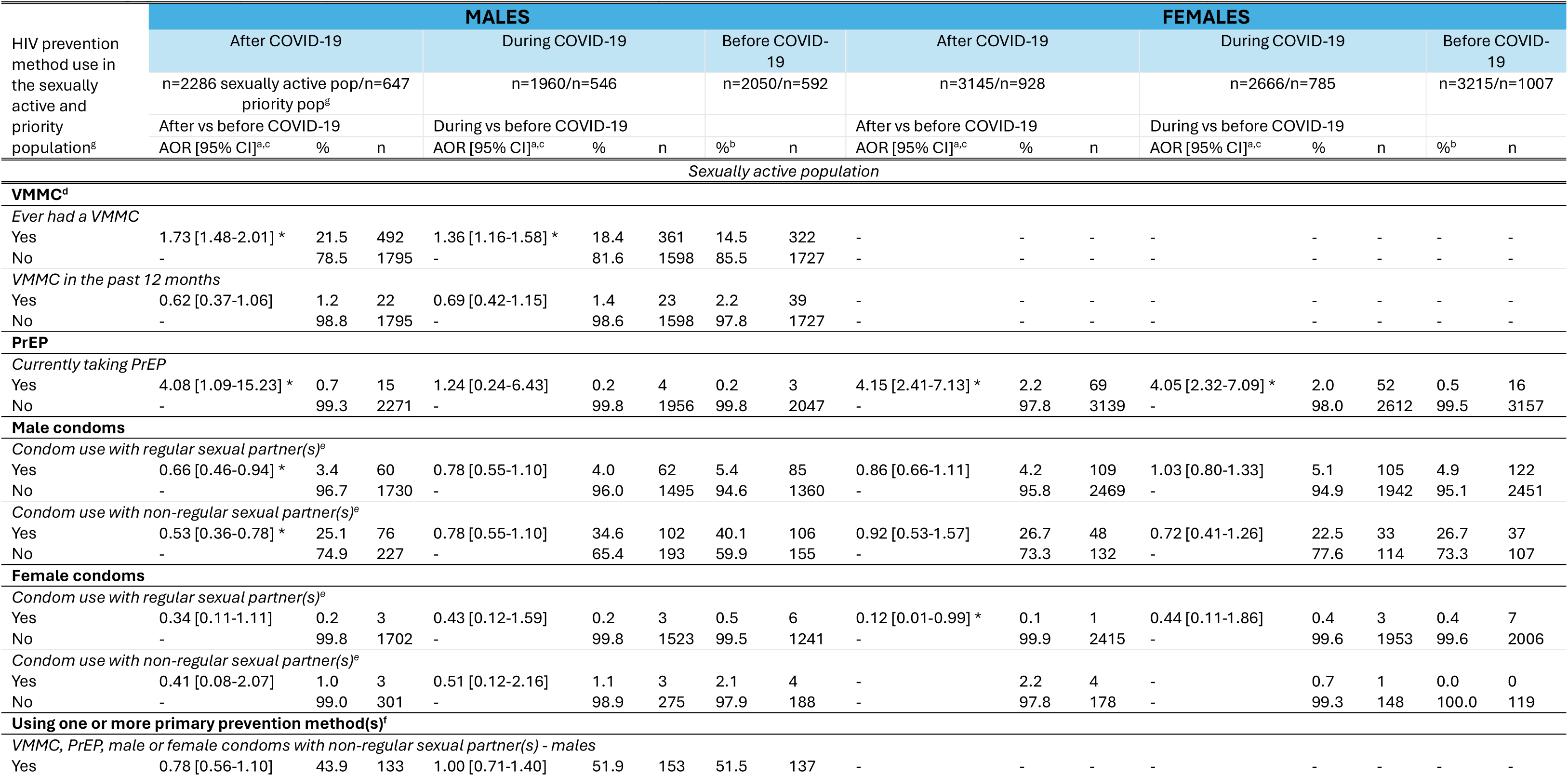
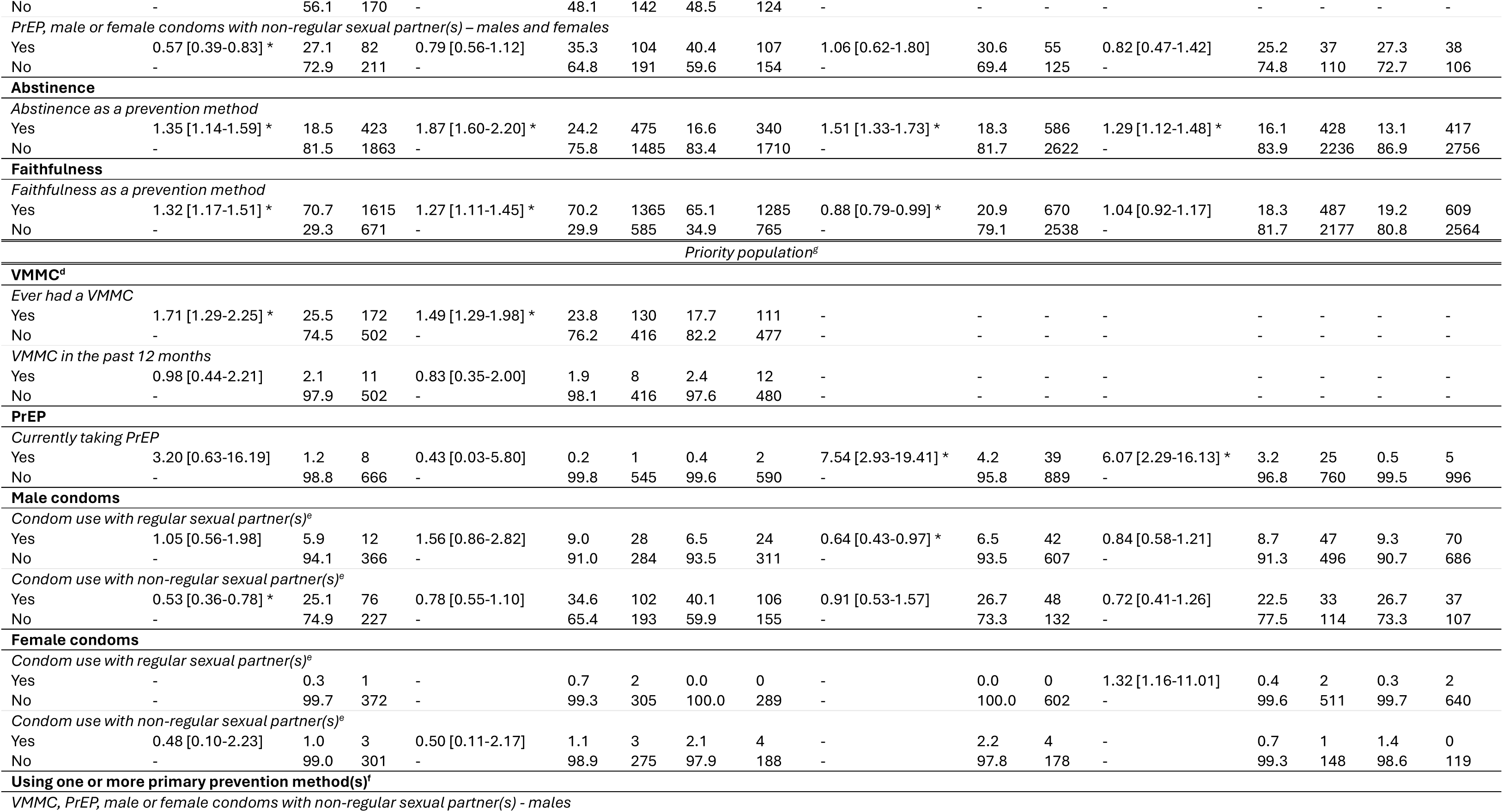
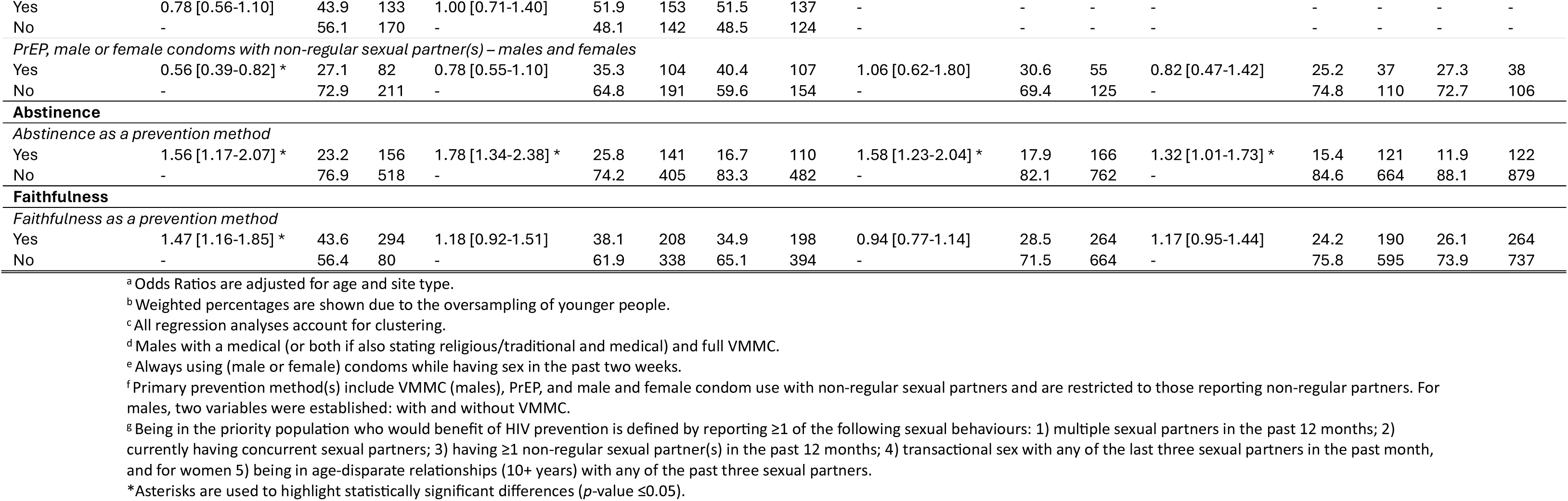
Comparisons of use of HIV prevention methods in sexually-active HIV-negative adults and in those at high risk of acquiring HIV infection (the priority population) aged 15-54 years in east Zimbabwe, after and during COVID-19 *versus* before COVID-19. Serial cross-sectional analysis.

For males, in the post-COVID-19 survey, the odds of consistently using condoms were lower for both regular [AOR=0.66, 0.46-0.94] and non-regular [AOR=0.53, 0.36-0.78] sexual partners. PrEP use was higher after COVID-19 [AOR=4.08, 1.09-15.23], although still low (1.2%). Within the cohort, fewer males reported condom use with non-regular sexual partners [AOR=0.24, 0.10-0.59] after COVID-19 compared to before COVID-19; however, more males reported condom use with their regular sexual partner(s) during COVID-19 [AOR=2.97, 1.16-7.58] (Figure 3e; [18]). Reports of sexual abstinence and faithfulness to protect against HIV infection increased in both analyses.

For females, no changes were found in male condom use, while female condom use was notably lower after COVID-19. In both analyses, PrEP use was higher during [serial cross-sectional: AOR=4.05, 2.32-7.09; cohort: AOR=9.05, 2.36-34.74] (Figures 3c & 3g) and after COVID-19 [serial cross-sectional: AOR=4.15, 2.41-7.13; cohort: AOR=6.91, 1.85-25.78] than before COVID-19 (Figure 3d & 3h). More women reported sexual abstinence during and after COVID-19 than before COVID-19.

VMMC was not included in the composite HIV prevention variable as it is a permanent method and, therefore, is less sensitive to short-term events. In the serial cross-sectional analysis, sexually-active men were more likely to report ever having had VMMC during [AOR=1.36, 1.16-1.58] (Figure 3a) and after [AOR=1.73, 1.48-2.01] (Figure 3b) COVID-19 than before COVID-19. More men in the cohort reported lifetime VMMC after COVID-19 than before COVID-19 [AOR=1.66, 1.18-2.35]. However, no changes across the three survey rounds were found in either analysis in reports of VMMC in the past 12 months (Table 5; Figure 3; [18]).

Findings for the priority population for HIV prevention methods were similar to those for sexually-active individuals [18]. Notably, for females, in the serial cross-sectional analysis, PrEP use increased quite substantially from 0.5% before COVID-19 to 3.2% during COVID-19 [AOR=6.07, 2.29-16.13] and 4.2% after COVID-19 [AOR=7.54, 2.93-19.41].

## 4. Discussion

We found notable reductions in use of HIV prevention methods among populations in need of these methods (i.e. those engaging in SRBs), and in key mediating social behaviours, in the general population in Manicaland, east Zimbabwe, during and following COVID-19. In an earlier report, we documented continuing high levels of male SRB in the same population during COVID-19 [2], and the new data provide evidence that these levels of HIV risk behaviour continued through to the end of the pandemic. Taken in combination, these are very worrying findings that suggest that COVID-19 may have led to an increase in new HIV infections [7].

### 4.1 Synthesis of findings

HIV prevention method use in Manicaland decreased for some methods and increased for other methods during COVID-19. Most notable is the decrease in males reporting condom use with non-regular partners. This echoes a finding in Kenya [19] and likely reflects, in part, disruptions in condom access that occurred in our study areas during COVID-19 [9] and have also been reported in slum areas in South Africa [8]. The scale-back in international funding for male condom programmes in Zimbabwe [20] may have contributed to the reduced access. The increase in females reporting PrEP use during and after COVID-19 is encouraging and may reflect low use before COVID-19, due to its recent introduction, and recent implementation of community-based provision [21].

During COVID-19, the serial cross-sectional data showed no overall changes in the proportions of men and women engaging in SRB in Manicaland [2]. However, the cohort analysis results revealed extensive individual-level changes. Large numbers of men (especially) and women at low risk of acquiring HIV infection prior to COVID-19 adopted higher-risk behaviours during COVID-19 but similar numbers of those at high risk initially adopted safer behaviours [2]. In the current study, this pattern of counteracting increases and decreases in different individuals’ HIV risk behaviour, resulting in no change in population-level SRB, was found to have recurred in the later stages of COVID-19.

Some changes in individuals’ SRB may have been life-course changes unrelated to COVID-19 but others could have stemmed from COVID-19-related shifts in personal circumstances [9]. In males, the only population-level change in SRB by the end of COVID-19 was a reduction in reports of concurrent sexual partnerships. This was despite reported reductions in beerhall visits – reflecting lockdown closures and restricted opening hours [13] – and use of recreational drugs; both social behaviours associated with SRBs in Manicaland [22]. In females, reports of multiple sexual partners and transactional sex declined; unexpected results, as increases occurred in earlier pandemics [23, 24]. A possible explanation for the different findings in males and females is that, whilst beer-hall closures reduced the numbers of women engaging in bar-based sex work, men may have formed other types of non-regular relationships with fewer women. In Uganda, some men had to end multiple transactional partnerships and strategically choose which ones to maintain [25]. In Manicaland, women’s reports of IPV are associated with their own and their marital partners’ SRB, as well as alcohol consumption [26]. Therefore, the reduction in IPV during COVID-19 – an unexpected finding despite being reported elsewhere [27] – might be explained by the beerhall closures and associated reductions in male partner alcohol consumption [28].

### 4.2 Strengths and limitations

Strengths of this study include its large sample sizes; diverse study sites; and unique combination of data on use of HIV prevention methods, SRBs, underlying social behaviours, and sociodemographic characteristics. Access to both serial cross-sectional and cohort data spanning COVID-19 enhanced inference and partly mitigated limitations associated with relying solely on one method, such as attrition bias in longitudinal data.

Limitations include difficulties in establishing a causal relationship with COVID-19 in the absence of a counterfactual scenario. Some SRBs were measured over 12-month periods that, during the COVID-19 surveys, spanned periods of varying restrictions. Changes in interview modality, from face-to-face interviews in round 7 (pre-COVID-19 baseline) to telephone interviews in subsequent rounds, may have resulted in changes between surveys in participation bias. Study participation rates in rural areas generally are higher in face-to-face interviews (due to more time spent at home) and lower in telephone interviews (because of gaps in network coverage); the opposite holds true for urban areas. Males can be easier to reach via telephone, as they often spend time away from home during the day, and older individuals may be more inclined to answer their phones while valuing phone calls [29]. In our population, we observed a slight increase in the mean age of participants between surveys. To address these issues, all analyses were stratified by sex and adjusted for age and site type. The changes in interview modality also may have caused changes in social desirability bias. Increased under-reporting of illicit substance use has been found in telephone interviews [30] and could also occur for other sensitive topics such as IPV and sexual relationships. We minimised these biases by training survey enumerators to be alert and pause interviews when others were present, and to build rapport, as well as by including selected questions multiple times to ensure greater intra-item validity [31].

Our finding of increases in lifetime VMMC over COVID-19 is implausible as the Zimbabwe Ministry of Health and Child Care (ZMOHCC) scaled down VMMC services in March 2020 [32] and VMMC procedures declined from 24,000 per month in early 2020 to a few hundred in April 2023 [3]. VMMC was probably over-reported in the telephone interview surveys conducted during and after COVID-19 as photographs could not be used to illustrate different forms of male circumcision. Consequently, we excluded it from the primary measure of combination HIV prevention reported in the study.

### 4.3 Preparing for the next pandemic: policy implications

Our findings add to growing evidence that the COVID-19 pandemic severely undermined the HIV response [10]. This is of great concern for two reasons. First, because – particularly in SSA where HIV infections remain at hyperendemic levels [33] – there is a high potential for substantial rebounds in new infections and deaths if HIV services are not sustained. Second, there is a strong likelihood that new pandemics will arise in the near future. A recent Lancet Commission noted that: “Another way to present our assessment of risk is that there is about a 50% chance that a new pandemic causing 25 million or more deaths will occur between now and 2050.” (p.1568) [34]. Therefore, making plans now to ensure that access to HIV and related SRH services is maintained throughout future pandemics must be an urgent priority.

Suggested actions for such a plan could include: (1) recognising HIV services as essential and allowing travel to access these services [4, 11]; (2) ensuring consistent production and timely delivery of these services, with regular stock monitoring and the implementation of innovative delivery methods [11], such as digital ordering and community drop-off points to minimise face-to-face contact; (3) including condoms and PrEP on the list of essential commodities during lockdown periods [12] and expanding their availability beyond beerhalls and clinics; (4) promoting home-based HIV self-testing to reduce clinic visits and support early detection; (5) enhancing promotion of HIV prevention, testing, and treatment through social media during lockdowns [12]; (6) responding to the specific needs of vulnerable groups [12], and (7) tracking changes in HIV infections and deaths.

## Data Availability

Given the sensitive nature of the data collected, including details about HIV status, treatment and sexual risk behaviour, the Manicaland Centre for Public Health does not make full analysis datasets publicly available. Nevertheless, summary datasets encompassing household and background sociodemographic individual questionnaire data from rounds 1 to 9 (1998 to 2023) are available for download via the Manicaland Centre website. Additionally, summary data on HIV incidence and mortality from rounds 1 to 6 (1998 to 2013), developed in collaboration with the ALPHA Network, can be accessed via the DataFirst Repository. For other quantitative data used in the Manicaland Centre for Public Health analyses, interested parties can request information by filling out a data access request form.

http://www.manicalandhivproject.org/data-access.html

https://www.datafirst.uct.ac.za/dataportal/index.php/catalog/ALPHA/about

http://www.manicalandhivproject.org/data-access.html

